# Dynamic Changes and Pronounced State Level Disparities in Controlled Substance Distribution by US Advanced Practice Providers From 2006 to 2023

**DOI:** 10.1101/2025.10.16.25337625

**Authors:** Julia L. Soares, Kenneth L. McCall, John J. Coleman, Brian J. Piper

## Abstract

**Objective:** To evaluate the changing pattern of the distribution of Schedule II and III opioids, stimulants, and barbiturates by advanced practice providers (APP; i.e., physician assistants and nurse practitioners) in the United States (US).

**Design:** Retrospective study.

**Sample:** Advanced practice providers from every US state (and DC) that directly dispense Schedule II/III drugs to patients.

**Procedures:** Controlled substances distributed by APP was obtained from the Drug Enforcement Administration’s Automated Reports and Consolidated Orders System (DEA’s ARCOS) for opioids (e.g., hydrocodone, fentanyl, buprenorphine), barbiturates (pentobarbital, butalbital), and stimulants (amphetamine, methylphenidate, lisdexamfetamine) from 2006–2023. Opioids were converted to their morphine milligram equivalents (MME), stimulants converted to daily doses, and barbiturates to kilograms. Opioid use by state in three selected years (2013, 2020, 2023) was further analyzed.

**Results:** The total weight of controlled substances as distributed exhibited both overall and drug-specific changes since 2006. Buprenorphine accounted for only a modest amount (0.6%) in 2013 but the preponderance (94.6%) of opioids distributed by MME in 2023 nationally. Examination of the opioid MME per state and corrected for population revealed the states with the highest reported use for 2013 (Nevada), 2020 (North Dakota), and 2023 (Maine). There has been an overall modest decline in stimulant (-97.9%) and barbiturate (-64.1%) since 2010.

**Conclusions and Clinical Relevance:** The use of Schedule II/III drugs as distributed to APP has fluctuated yet overall increased in the past decades. Opioids by total MME have had rather small changes throughout the years, except for buprenorphine. APP direct distribution has transitioned from treating pain to Opioid Use Disorder. Future research should discover the causes underlying the yearly and state level disparities for opioid, stimulant, and barbiturate use.

## INTRODUCTION

There has been a prominent opioid epidemic spreading across the United States^1^ in recent decades which is linked to increased prescription of opioids for pain and other medical concerns.^2^ However, the CDC found a 27% decrease in drug overdose deaths in 2024 compared to the previous year.^3^ The medical specialties that prescribe have expanded to include advanced practice providers (APP, i.e., physician assistants and nurse practitioners—PAs and NPs). Due to this, APP had appreciable increases in their opioid prescriptions.^2^ PAs and NPs in almost every state can prescribe Schedule II drugs (i.e., opioids like hydrocodone and fentanyl) (**Supplemental Figure 1**) but less is known for the distribution for APPs.

A recent report examined overprescribing as defined by both how many patients they distributed the substances to (>50%) and how much of the substance (≥100 MME/day to more than 10% of their patients). PAs and NPs had have the highest rates of overprescribing opioids to their patients (9.8% and 8.0%, respectively).^21^ In addition, in states where APP were allowed to distribute opioids independently, they were over twenty times more likely to do so than in states where they faced restrictions.^21^ Overprescription of opioids, especially if they are not necessary to treat pain, plays a large role in maintaining or creating opioid addiction.^22^

Between 2003 and 2010, there was a stark increase of “pill mills” in Florida, where providers had been directly distributing large amounts of opioids and other prescriptions to patients in inappropriate or illegal ways to gain higher profits.^5^ Researchers found that this malpractice was contributing to the development of opioid use disorder (OUD) and deaths from overdose.^6^ In 2010, the Florida Department of Health (DOH) began to crack down on this issue and instated laws restricting direct prescriptions which helped to decrease the mortality rate of deaths from overdose^5^ and the prescriptions and use of opioids.^7^ Researchers have found that in Florida, the average amount of opioids distributed by providers increased by 120.6% from 2006 to 2010.^37^ They then found a decline in the distribution after 2010, after the Florida DOH laws were enacted.^37^ As news reports and data on these pill mills have focused on opioid prescriptions from providers, the question now stands of whether PAs and NPs have followed in the same footsteps in their prescription patterns during this time.

APP have taken on an important role in treating opioid use disorder, especially in their distribution of buprenorphine to patients.^8^ In 2016, NPs were given authority to prescribe buprenorphine to treat OUD to help fill the treatment gap for this population and are required to complete a twenty-four hour training course in order to prescribe the substance.^8^ Since the *Comprehensive Addiction and Recovery Act* was enacted, NPs and PAs made up more than half of the 111% increase in waivered clinicians in rural areas.^9^ Buprenorphine is regarded as a “gold standard” in OUD treatment and is associated with high decreases in mortality rates among users.^10^ It is also a first line of treatment for the disorder, as it can be prescribed and dispensed to patients directly from their provider.^18^ Buprenorphine acceptance and retention have been increasing across the United States.^11^ Although originally used only to treat pain, it has now been more commonly used for the treatment of OUD, including in combination with pain.^12^ NPs in fourteen states, particularly in the Northwest (AK, AZ, DC, HI, IA, ID, MT, ND, NH, NM, OR, RI, WA, and WY) in 2018 had full, autonomous practice and prescription authority without a requirement for physician supervision or collaboration.^16^ NPs residing in states with a requirement for physician oversight were over forty percent less likely to prescribe buprenorphine.^17^ The “X-waiver” to prescribe buprenorphine for OUD was eliminated in 2022 although individual states have their own requirements.

This paper aimed to analyze and evaluate the amount and registrants of Schedule II and III drugs (i.e., opioids, stimulants, and barbiturates) that were distributed to APPs and reveal recent distribution patterns by year and state.

## MATERIALS AND METHODS

### Procedure

Manufacturers and distributors annually report the distribution of controlled substances to the Drug Enforcement Administration’s (DEA) Diversion Control Division. Controlled substances distributed by APP (referred to by the DEA as “mid-level providers”) was obtained from report seven of the DEA’s Automated Reports and Consolidated Orders System (ARCOS) for opioids (e.g., hydrocodone, fentanyl, codeine), barbiturates (pentobarbital, butalbital), and stimulants (amphetamine, methylphenidate, lisdexamfetamine) from 2006–2023.^23^ There has been much controversy surrounding the most appropriate title for categorizing PAs and NPs.^13-15^ The US DEA typically refers to these providers as “mid-level providers” as to differentiate them from physicians (MD, DO) who are designated as “providers.”^22^ However, NPs and PAs have found this term to be confusing and belittling of their important medical work.^4^ Due to this, we have elected to refer to this group of practitioners as APP, which is a more accepted and commonly utilized term. Procedures were approved by the IRB of Geisinger.

### Data-analysis

Opioids were converted to their morphine milligram equivalents (MME). This was calculated by multiplying the weight in grams by the opioid’s conversion factor: buprenorphine: 10, codeine: 0.15, fentanyl: 75, hydrocodone: 1, hydromorphone: 4, meperidine: 0.1, methadone: 8, morphine: 1, oxycodone: 1.5, oxymorphone: 3, and tapentadol: 0.4.^19^ Next, stimulants were converted to daily doses: methylphenidate and amphetamine have a daily dose conversion of 20 mg/day/person, and lisdexamfetamine’s daily dose is 40 mg/day/person.^20^ In order to calculate this, weights were converted from grams to milligrams and then divided by 20 or 40, respectfully. Barbiturate use was converted from grams to kilograms. Pentobarbital data was primarily focused on in the calculations, as the data for butalbital and barbituric acid derivative/salt was negligible. The drugs with the highest use were analyzed for patterns over time.

In addition, opioid use by state as distributed to APPs was obtained for 2013, 2020, and 2023. The years 2013 and 2020 were the peak years for opioid distribution, so we chose to expand upon the specific drug weights in those years, as well as including use and registrant data for 2023 as the most recent (when data was collected in 8/25) full-year for comparison. State populations for those three years were collected from US Census records for calculations of MME per 10,000 persons. Figures were prepared with GraphPad Prism (Version 10.5.0, San Diego, CA).

## RESULTS

The total weight of controlled substances distributed directly by APP exhibited both overall and drug-specific change since 2006. Across the US, the total weight of opioids in MME peaked in 2023 (**Supplemental Figure 3A**). Hydrocodone peaked in 2012 and fentanyl in 2017, while the total use of opioids is continuing to increase as of 2023, particularly buprenorphine.

Figure 1. reveals that hydrocodone accounted for a plurality (45.0%) of the opioids distributed by APP in 2013. Similarly, buprenorphine accounted for a negligible (0.08%) portion of all opioids by MME in 2006 (6.8) versus the preponderance (94.6%, 60,762.7 MME) in 2023. Fentanyl was shown to be the highest drug distributed by providers in 2013 (33.5%), while in later years hydromorphone (2020, 44.6%) and buprenorphine (2023, 65.1%) rose to the top. In 2013 and 2020, pharmacies distributed oxycodone the most compared to the other opioids (31.8%, 30.8%), while buprenorphine accounted for the largest amount in 2023 (38.8%). Methadone distribution by pharmacies has also shown a decrease over the years from 18.9% in 2013 to 7.4% in 2023.

**Figure 1.**
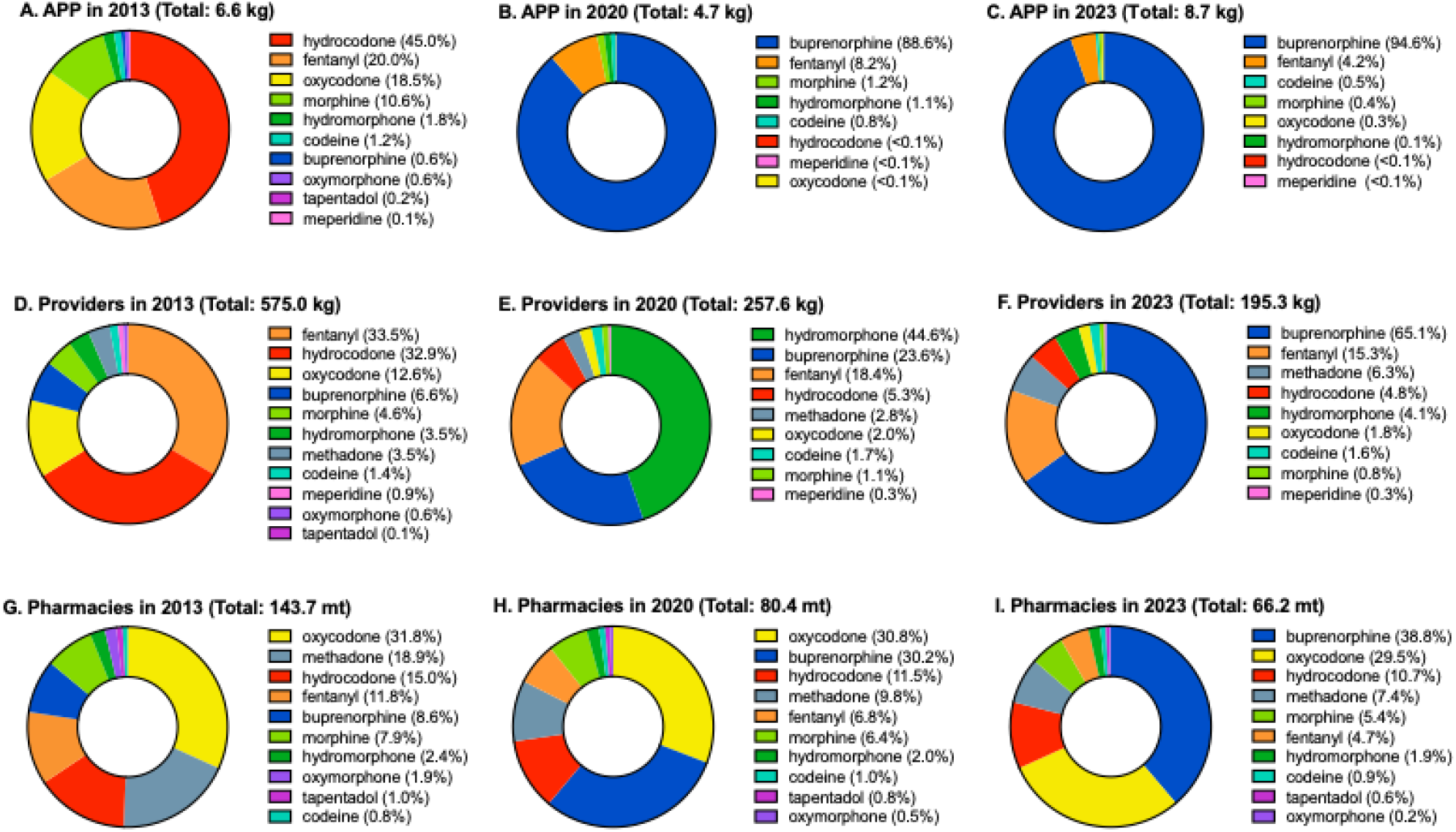
Total weight of prescription opioids (i.e., buprenorphine, codeine, fentanyl, hydrocodone, hydromorphone, meperidine, methadone, morphine, oxycodone, oxymorphone, tapentadol) by morphine mg equivalents (MME) as distributed by Advanced Practice Providers, Providers, and Pharmacies as reported to the Drug Enforcement Administration’s Automated Reports and Consolidated Orders System (ARCOS) in 2013 (A, D, G), 2020 (B, E, H), and 2023 (C, F, I).^23^

There were also stark differences in the distribution patterns between other practitioners and APP regarding the total number of registrants and total MME for the Schedule II and III substances across the country. For example, in 2023 practitioners distributed 438,218.0 MME of buprenorphine to 30,442 registrants total, while APP prescribed 60,762.7 MME to 290 buyers overall.^23^ In 2023 there were reported to be roughly 280,140 NPs and 145,740 PAs nationwide, while there were about 1,010,892 active physicians (851,282 in direct patient care).^26-27^ Furthermore, in 2023, there were 6,129 hospitals across the nation and 40,194 pharmacies.^28-29^ In 2023, pharmacies distributed roughly 53,242,925.5 MME of buprenorphine to 57,899 buyers total, while hospitals prescribed 2,834,195.0 MME to 6,822 total registrants.^23^

The number of registrants for opioids have shown an overall decrease since 2006, with hydrocodone purchases peaking at around 3,000 buyers in 2012, fentanyl at 1,500 buyers in 2017. However, the number of fentanyl registrants has seen a slight increase in 2023. The amount of buyers of buprenorphine was modest in 2006 (7) but increased over forty-fold to 290 in 2023 (**Supplemental Figure 2B**).

Figure 2. shows that the states with the highest use of opioids per population (MME/State Pop x 10,000) have changed over the past decade. Nevada reported the highest use per population rate of 26.2, while Alaska (2.9) and Alabama (2.2) were the next highest states in 2013. North Dakota was the top distributor of opioids (16.0) with Maine (11.0) and Alaska (7.0) following closely behind in 2020. Maine had the highest distribution (80.3) followed by Alaska (15.7) and Oregon (12.5) in 2023. The patterns from these three years show a slight decrease and then substantial increase, yet an overall fluctuation in the weight of opioids being distributed by APP over time per state. The distribution of opioids was very non-homogenous across the US with the top five states accounting for 83.4% of the total MME in 2013, 70.3% in 2020, and 72.5% in 2023.

Further analysis was completed on the highest distributing states. **Supplemental Figure 5** shows that oxycodone accounted for 46.1% of the total MME in Nevada in 2013, and buprenorphine accounted for 99.0% in North Dakota in 2020 and 99.9% in Maine in 2023.

**Supplemental Figure 3** reveals that the number of stimulant buyers was negligible.

**Figure 2.**
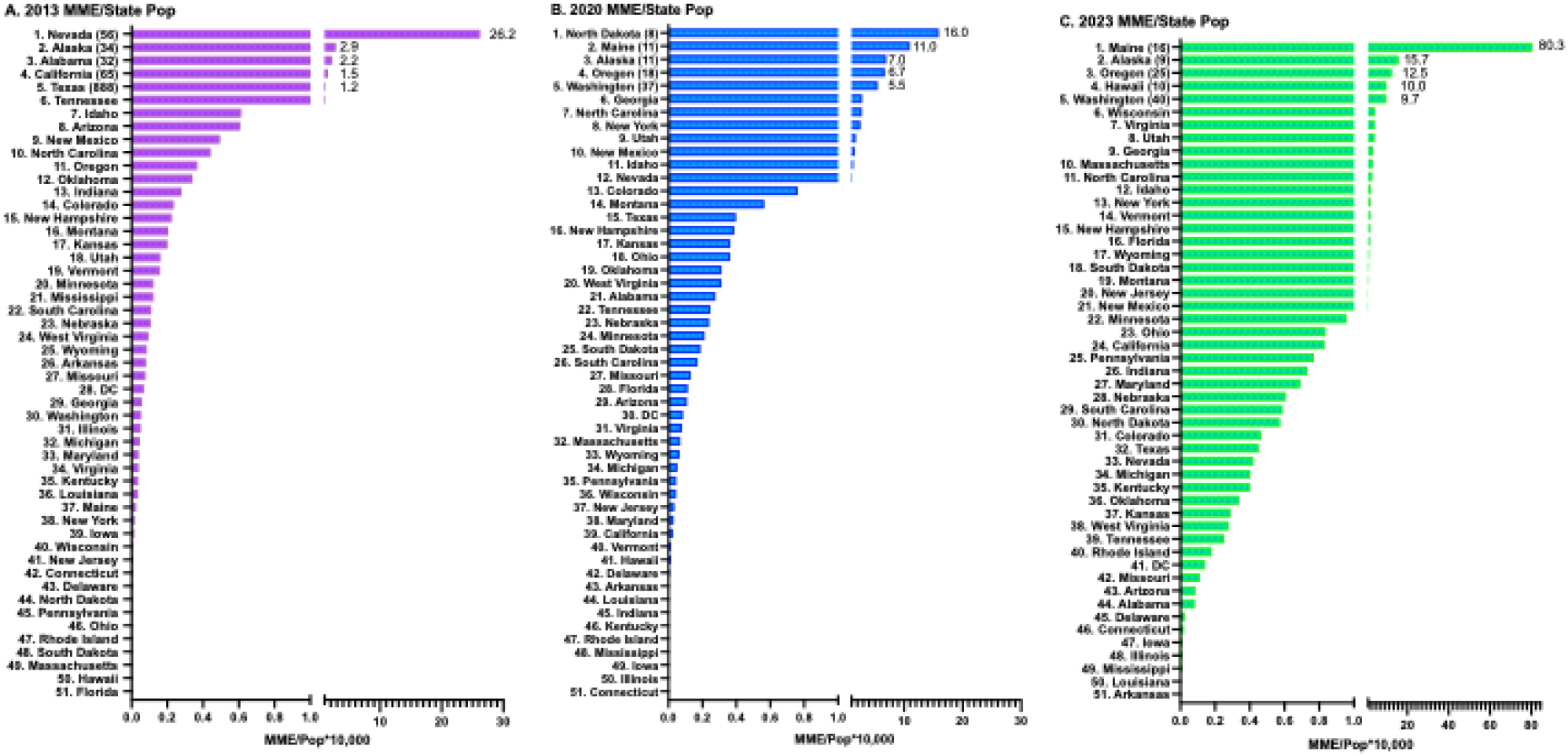
Total weight in morphine mg equivalents of opioids per each person of each US state in 2013, 2020, and 2023 distributed by Advanced Practice Providers as reported to the Drug Enforcement Administration’s Automated Reports and Consolidated Orders System (ARCOS).^23^ The number of registrants is shown in parenthesis for each of the top five states.

**Supplemental Figure 4** shows that the number of registrants for barbiturates, pentobarbital in particular, peaked in 2011, with a steady decline until 2016. The total weight of barbiturates followed the same pattern as pentobarbital with a peak in 2007 and decreased until 2016, with a slight peak in use in 2010. Note that pentobarbital has not been reported since 2016.

## DISCUSSION

This novel investigation determined that use of Schedule II and III drugs as distributed by APP directly to patients has fluctuated in the recent years but is in an overall upward trajectory for opioids and downward for stimulants and barbiturates. Across states, the distribution of opioids was non-homogenous, which could at least partially be attributed to the state differences in prescribing power for PAs and NPs^16-17^ (**Supplemental Figure 1**). Reported opioid use and buyers were on the decline from 2006-2010, then increased and peaked in 2012-2013. Opioid MME per state population analyses revealed the states with the highest reported use for 2013 (Nevada), 2020 (North Dakota), and 2023 (Maine). The number of registrants steadily increased for fentanyl, peaking in 2017, while the overall use of all reported opioids has been continuing to rise since 2018. Data on amphetamine use and buyers showed a noticeable uptick between 2009 and 2015, with a small increase in registrants in 2019, but an overall decrease in reported use. The number of registrants for barbiturates peaked in 2011 and steadily decreased, while the use of this drug class fluctuated between 2006-2010 before reporting a downward pattern as well.

As of 2024, state laws have shown some variation in condoning physician assistants (PA) and nurse practitioners (NP) to prescribe schedule II drugs (**Supplemental Figure 2**). Physician assistants were forbidden from prescribing schedule II substances in Alabama, Arkansas, Georgia, Hawaii, Iowa, and West Virginia. Nurse practitioners were not allowed to prescribe this category of agents in Georgia, Oklahoma, South Carolina, and West Virginia. While not fully restricted, PAs and NPs in Arkansas and Missouri are only allowed to prescribe hydrocodone products from the list of schedule II substances.^30^ It is interesting to note the differences on state policies for two types of the same level of provider, in addition to the pattern of southern/central states being more restrictive in the prescriptive actions of APP. Relative to prescribing privileges, this data indicates that direct dispensing by APP of opioids and barbiturates is very modest in the majority of states and that dispensing of stimulants is exceedingly uncommon.

In examining the trends of NP’s and PA’s scope-of-practice regulations by state from 2001 to 2010, researchers found that many states across the country loosened their regulations allowing APP to hold greater prescriptive authority with less involvement from physicians.^31^ Due to these changes in regulations, there have been positive effects for patients, such as reduced outpatient care costs while still maintaining the same level of care intensity.^32^ After a state reduces their restrictions on APP prescriptive authority, the overall net amount of opioids decreases.^33^ This shows that although the amount of opioids dispensed by APP increases slightly, the decrease in physician prescriptions create an overall decline.^33^

While buprenorphine was originally created in the 1960s to be used as a pain reliever that could be used in place of morphine, it is now more commonly used to treat OUD.^34^ As we have found that the distribution of all opioids by APP has seen an increase in the recent years, buprenorphine has made up a large preponderance of that. This shows that although the use of other opioids for pain, such as fentanyl and hydrocodone has not increased, the treatment for addiction to opioids by APP has risen greatly. Furthermore, with regard to the provider-led “pill mills” discussed earlier^5^, we have found that PAs and NPs increased their distribution of buprenorphine over the years to treat OUD in their patients, rather than increase the growth of the opioid epidemic. This shows that they have been working to decrease overdose and addiction related to opioid use and abuse and save lives in the process.

Strengths of this research include the availability of longitudinal, retrospective data that spans across different forms of substances, states, and providers. This comprehensive data set allowed us to analyze the patterns of buyers and amounts of drugs distributed by APP since 2006. In addition, in using preexisting data, we were able to focus on the analysis of the data instead of its collection. Although prior studies have examined direct distribution of Schedule II opioids by providers,^35-36^ including excesses associated with “pill mills” in Florida,^37^ this is the first report focusing on APP.

There are some limitations and caveats with ARCOS. First, although there are roughly 189,907 PAs^38^ and 385,000 NPs^39^ in the US, further study with other national and comprehensive databases will be necessary to characterize each of their controlled substance prescribing patterns directly to patients. Second, the DEA does not typically release information by National Drug Code so it is not possible to examine formulations (e.g. buprenorphine monoproduct versus buprenorphine with naloxone). include using pre-collected datasets from 2006 to 2023, in which the drugs reported varied slightly by the year—certain drugs were added in later in the years and others ceased to be reported at all. This led to dynamic changes in the data, such as pentobarbital’s reported use and buyers past 2016. Future research should be done on this topic to discover the causes behind the yearly and state trends for opioid, barbiturate, and stimulant use as distributed by APP in the United States. In addition, similar data on opioid distribution by providers (e.g., MD/DO)^35-37^ should be compared to this dataset and analyzed for similarities and differences.

### Conclusion

This paper analyzed and evaluated the amount (e.g., weight, daily dose) and number of registrants of Schedule II and III drugs (i.e., opioids, stimulants, and barbiturates) that were distributed by Physician Assistants and Nurse Practitioners and examined recent distribution patterns by year and state. We found that there has been an increase in opioid use, especially buprenorphine, and a decrease in stimulant and barbiturate use across the nation. We have also found state-specific differences in the distributions, as there are a small number of states in which APP are not allowed to distribute Schedule II substances. State distributions have also varied by substance, as certain states have higher usage of specific substances (e.g., hydrocodone, buprenorphine) than others, which has also changed overtime. This paper adds to the growing amount of research conducted on PA and NP prescribing patterns, particularly their patterns over the past two decades (2006-2023) and by state.

## Supporting information

Raw Data Set

## Data Availability

All data produced are available online at https://www.deadiversion.usdoj.gov/arcos/retail_drug_summary/arcos-drug-summary-reports.html

---

**Supplementary Figure 1.**
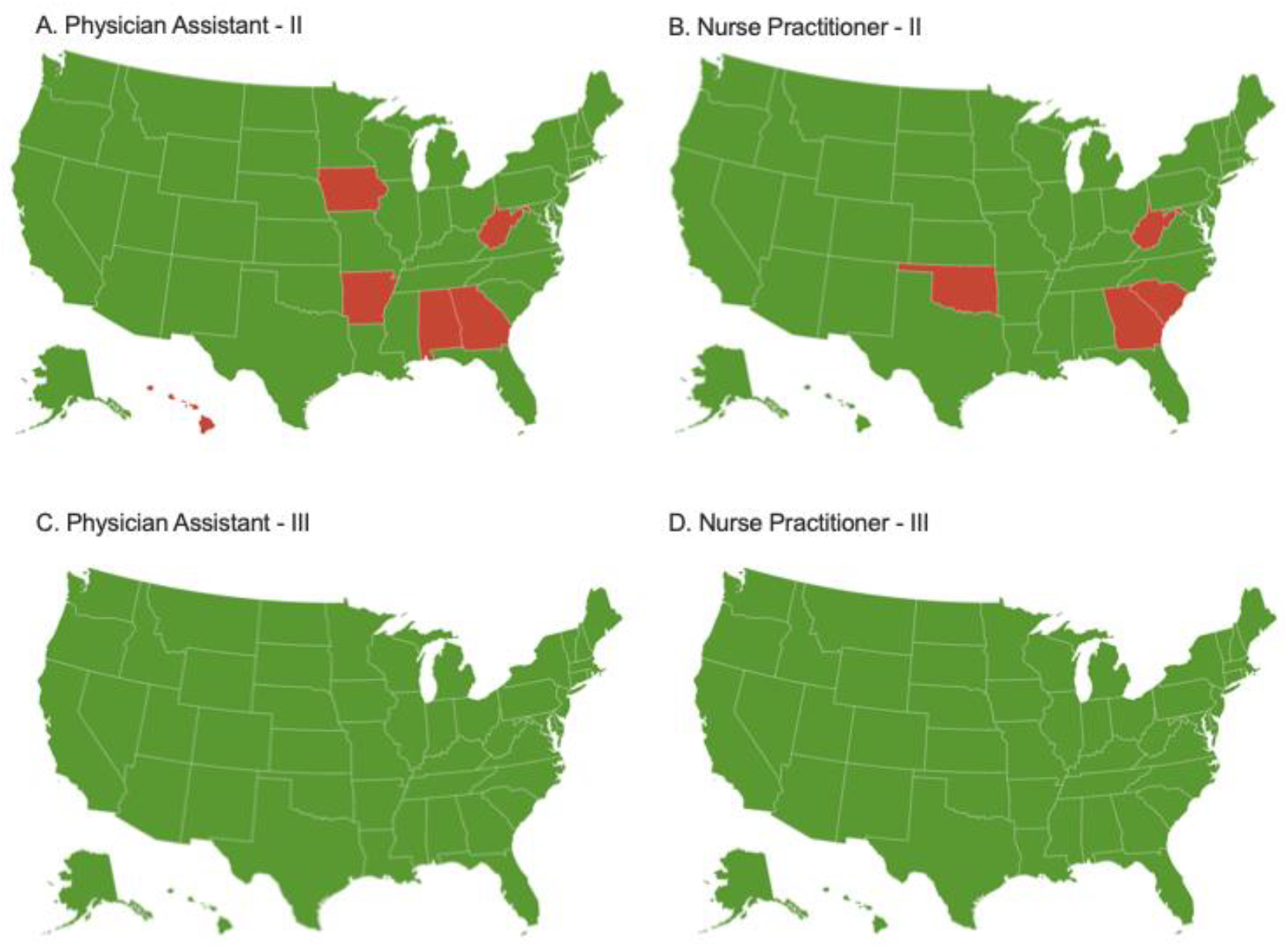
State laws in 2024 regarding Physician Assistants (A) and Nurse Practitioners (B) authority to prescribe Schedule II controlled substances (green = approved, red = prohibited), and regarding PA (C) and NP (D) authority to prescribe Schedule III controlled substances (green = approved). Information regarding the state laws of whether physician assistants and nurse practitioners are allowed or prohibited from prescribing schedule II and III drugs was obtained from the American Medical Association’s Advocacy Resource Center.^24-25^

**Supplementary Figure 2.**
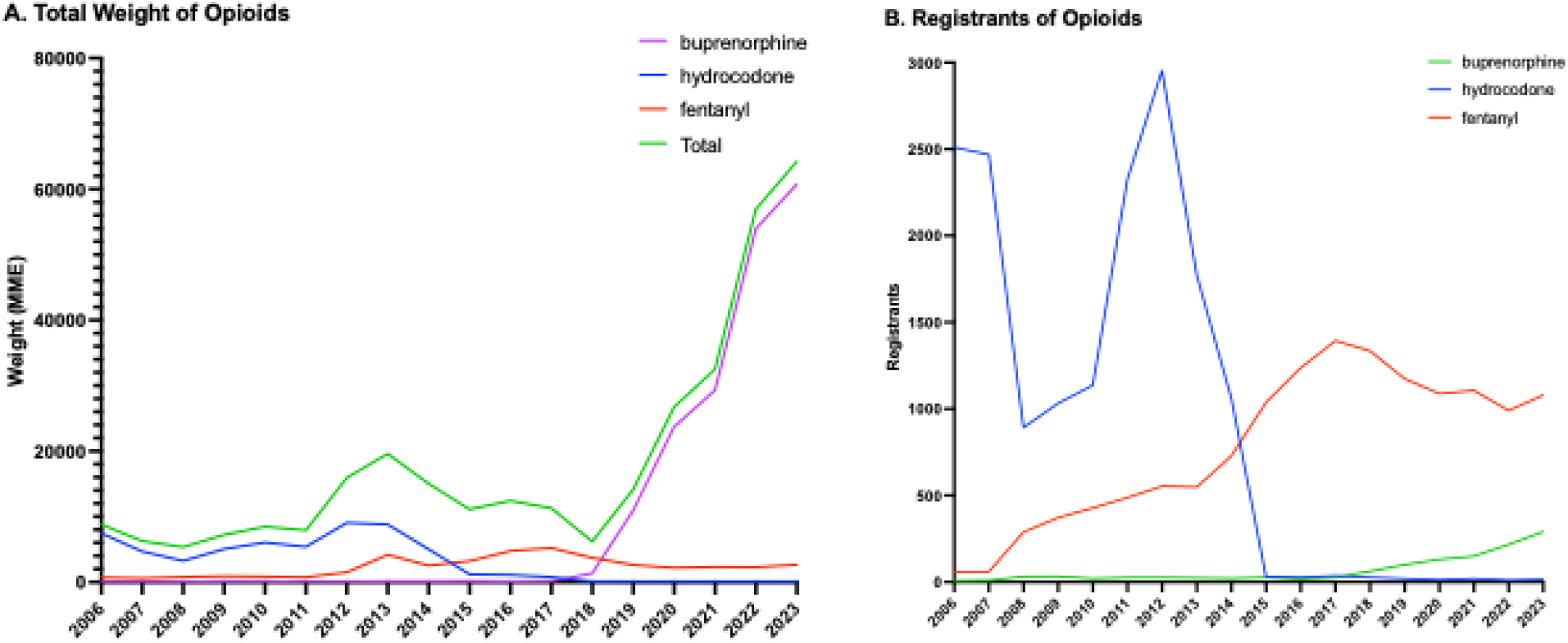
Total weight (A) of prescription opioids (i.e., buprenorphine, codeine, fentanyl base, hydrocodone, hydromorphone, meperidine, morphine, oxycodone, oxymorphone, tapentadol) by morphine mg equivalents (MME) and registrants (B) as distributed by advanced practice providers, 2006-2023 as reported by the Drug Enforcement Administration’s Automated Reports and Consolidated Orders System (ARCOS).^23^

**Supplementary Figure 3.**
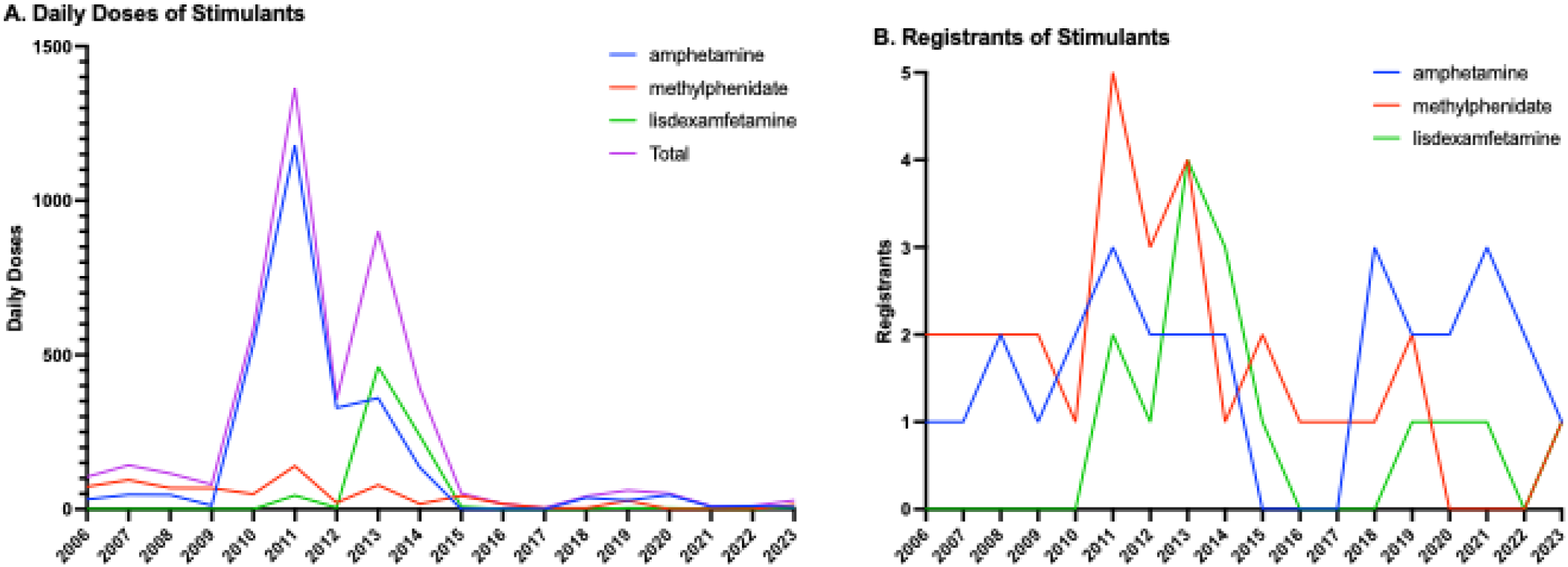
Registrants and daily doses of stimulants^5^ as distributed by Advanced Practice Providers, 2006-2023 as reported by the Drug Enforcement Administration’s Automated Reports and Consolidated Orders System (ARCOS).^23^

**Supplementary Figure 4.**
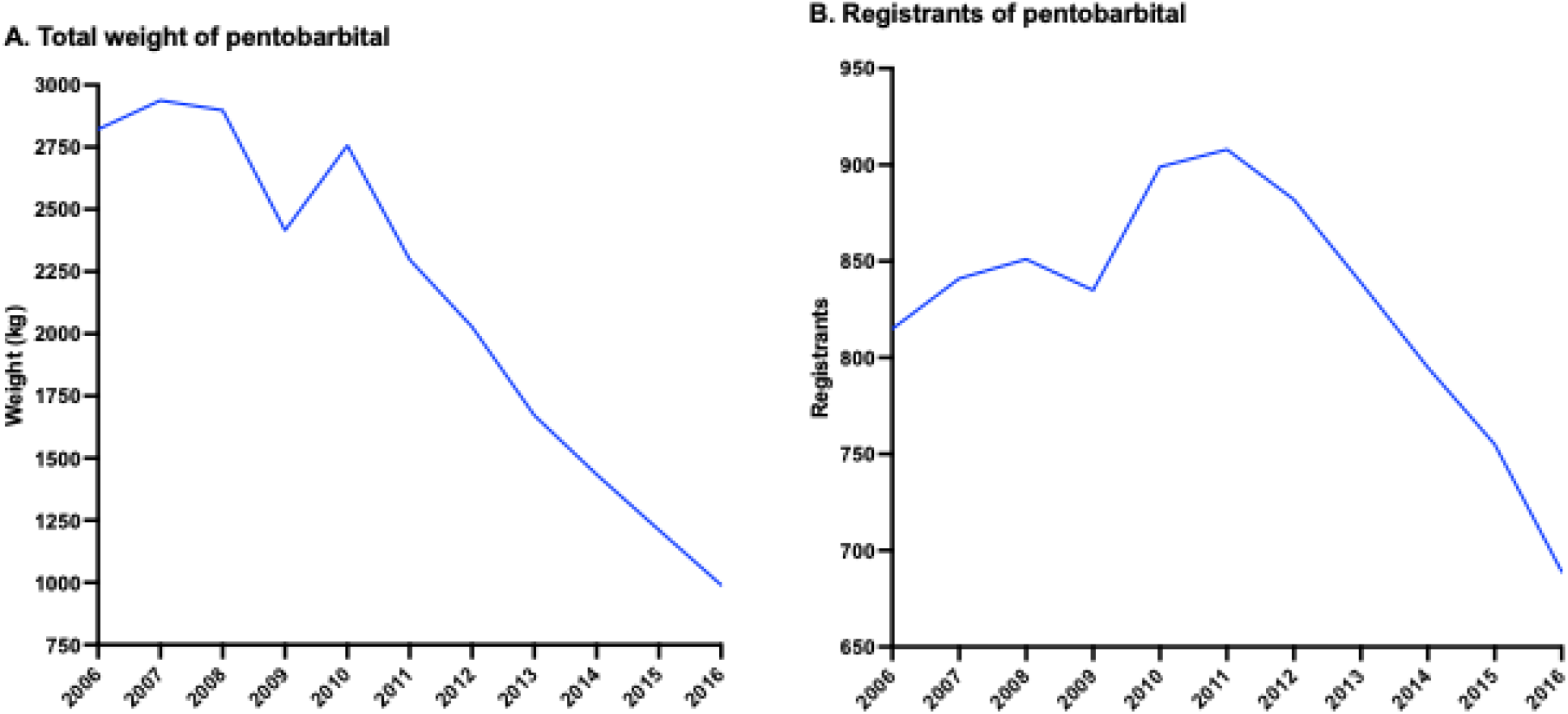
Registrants and total weight of pentobarbital as distributed by Advanced Practice Providers, 2006-2016 as reported by the Drug Enforcement Administration’s Automated Reports and Consolidated Orders System (ARCOS).^23^ Barbituric acid derivative or salt and butalbital had negligible weight (< 450g) and registrants (< 8 per year) and were not shown.

**Supplementary Figure 5.**
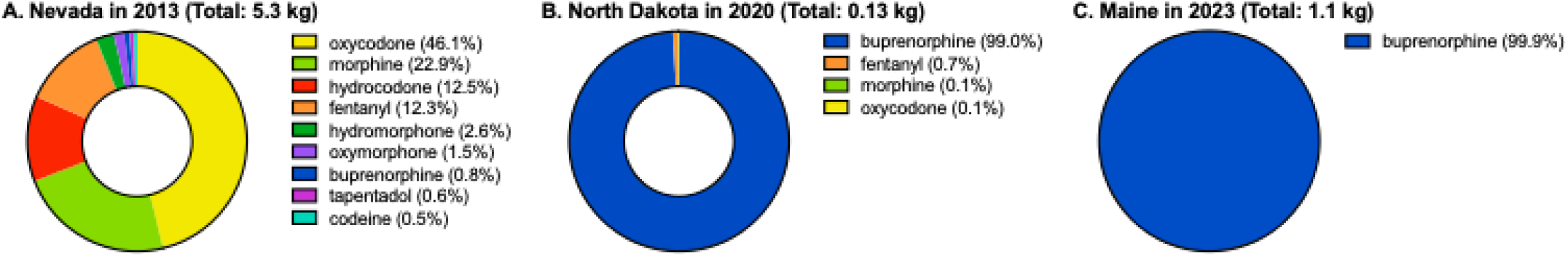
Total weight of prescription opioids (i.e., buprenorphine, codeine, fentanyl, hydrocodone, hydromorphone, meperidine, morphine, oxycodone, oxymorphone, tapentadol) by morphine mg equivalents (MME) as distributed by Advanced Practice Providers as reported to the Drug Enforcement Administration’s Automated Reports and Consolidated Orders System (ARCOS) in Nevada in 2013 (A), North Dakota in 2020 (B), and Maine in 2023 (C).^23^

